# Vitamin D for glycemic control following an acute pulmonary exacerbation: A secondary analysis of a multicenter, double-blind, randomized, placebo-controlled trial in adults with cystic fibrosis

**DOI:** 10.1101/2024.01.04.24300862

**Authors:** Alisa K. Sivapiromrat, William R. Hunt, Jessica A. Alvarez, Thomas R. Ziegler, Vin Tangpricha

## Abstract

Individuals with cystic fibrosis (CF) often incur damage to pancreatic tissue due to a dysfunctional cystic fibrosis transmembrane conductance regulator (CFTR) protein, leading to altered chloride transport on epithelial surfaces and subsequent development of cystic fibrosis-related diabetes (CFRD). Vitamin D deficiency has been associated with the development of CFRD. This was a secondary analysis of a multicenter, double-blind, randomized, placebo-controlled study in adults with CF hospitalized for an acute pulmonary exacerbation (APE), known as the Vitamin D for the Immune System in Cystic Fibrosis (DISC) trial (NCT01426256). This was a pre-planned secondary analysis to examine if a high-dose bolus of cholecalciferol (vitamin D_3_) can mitigate declined glucose tolerance commonly associated with an acute pulmonary exacerbation (APE). Glycemic control was assessed by hemoglobin A1c (HbA1c) and fasting blood glucose levels before and 12 months after the study intervention. Within 72 hours of hospital admission, participants were randomly assigned to a single dose of oral vitamin D_3_ (250,000 IU) or placebo, and subsequently, received 50,000 IU of vitamin D_3_ or placebo every other week, beginning at month 3 and ending on month 12. Forty-nine of the 91 participants in the parent study were eligible for the secondary analysis. There were no differences in 12-month changes in HbA1c or fasting blood glucose in participants randomized to vitamin D or placebo. A high-dose bolus of vitamin D_3_ followed by maintenance vitamin D_3_ supplementation did not improve glycemic control in patients with CF after an APE.

## 1. Introduction

Cystic fibrosis-related diabetes (CFRD) is the most common non-pulmonary co-morbidity associated with cystic fibrosis (CF) with a prevalence of up to 40-50% of adults with CF [1]. CF is a multi-organ disease caused by altered chloride transport on epithelial surfaces, leading to thick mucus secretions in organs such as the lung and pancreas, resulting in chronic infection, inflammation, and damage [2, 3]. Thickened mucosal secretions in the pancreatic ducts can obstruct and damage pancreatic tissue, leading to CFRD [1, 4]. Destruction of exocrine pancreatic tissue leads to fat malabsorption, resulting in undernutrition and vitamin deficiencies, including fat-soluble vitamins like D [5–7, 8]. Destruction of endocrine pancreas tissue contributes to disturbances in insulin secretion, and ultimately, CFRD in some individuals [1, 4].

Before CFRD onset, patients with CF experience significant pulmonary status and lung function decline [9], and consequently, a higher risk for recurrent pulmonary exacerbations [10]; however, insulin therapy has been seen to mitigate some of the negative effects [11]. Similarly, it has been suggested that untreated hyperglycemia has been associated with declined recovery from acute pulmonary exacerbation (APE) [12]. Hyperglycemia increases glucose within the airways, which allows bacteria to replicate faster, possibly contributing to an increased frequency of pulmonary exacerbations and worsened recovery [13, 14]. Continuous glucose monitoring in a patient with CF showed significantly higher interstitial glucose values in the week prior to a pulmonary exacerbation, suggesting APE frequency and glucose intolerance may be associated [14]. Therefore, improving glycemic control in patients with CF may enhance quality of life and lower mortality.

Vitamin D deficiency has been associated with prediabetes, insulin resistance, and an increased likelihood of developing diabetes [15–22]. Vitamin D deficiency is seen to have a causal effect on less efficient intestinal absorption of calcium [23]. Adequate calcium absorption may be beneficial to decrease the risk of CFRD due to its ability to stimulate insulin release to the blood and its possible indirect effect on weight loss [24–27]. Further, in the context of CF, vitamin D_3_ (cholecalciferol) supplementation has been associated with improved insulin secretion and decreased risk for CFRD [8, 28]. In a systematic review of seven randomized control trials of children with obesity, four trials indicated that vitamin D supplementation had a positive impact on glycemic control as determined by insulin levels, insulin resistance, and fasting blood glucose [29]. Prior studies have indicated differing results, especially about optimal vitamin D dosage and duration. A recent study in rats with type 2 diabetes (induced by a high carbohydrate and fat diet) indicated vitamin D_3_ treatment can counteract changes induced by hyperglycemia, therefore, diminishing metabolic dysregulation seen in the diabetic state [30]. Consequently, vitamin D supplementation may improve glycemic control in patients with CF.

Following critical illness or injury, such as an APE, blood glucose levels rise in response to metabolic effects, such as increased glucocorticoid and catecholamine hormone concentrations as well as greater insulin resistance in peripheral tissues [31–33]. The potential mechanisms for this acute development of insulin resistance are increased levels of suppressors of cytokine signaling proteins, leading to less insulin receptor substrate signaling or increased circulating free fatty acids [33]. Studies showed that hyperglycemia is highly prevalent during pulmonary exacerbations in patients with CF [34]. Therefore, we proposed that vitamin D may have some therapeutic effects to curb hyperglycemia and insulin resistance following an APE in adults with CF.

To our knowledge, there have been no prior studies currently that have examined changes in glycemic control after treatment of a high-dose bolus of vitamin D_3_ supplementation following an APE. Further, very few studies examine hyperglycemia in CF patients after an APE. A study showed pediatric patients who were admitted for a pulmonary exacerbation of CF had hyperglycemia that seemed to improve upon exacerbation treatment initially but had declined glucose tolerance after extended follow-up (3.8 - 6.3 months) [34]. Studies have indicated that poor glycemic control in adults and children with CF is associated with worsened FEV1 recovery, suggesting untreated hyperglycemia can negatively affect recovery from APEs [12, 34, 35]. Hyperglycemia has been described as a potential adjustable factor to improve recovery from pulmonary exacerbations [34].

This study aimed to examine changes in glycemic control, quantified by changes in hemoglobin A1c (HbA1c) and fasting blood glucose values, after administration of a high-dose bolus of vitamin D in adults with CF after an APE. This study was a pre-planned secondary analysis of the Vitamin D in the Immune System in Cystic Fibrosis (DISC) study, which was a multicenter, double-blind, randomized, placebo-controlled clinical trial in which adults with CF were hospitalized for APE therapy and concurrently treated with a single high-dose vitamin D bolus followed by maintenance supplementation of vitamin D_3_ [36, 37]. We hypothesized that vitamin D supplementation at the time of an APE for CF would improve glycemic control or mitigate hyperglycemia and insulin resistance that is associated with critical illness.

## 2. Materials and Methods

### 2.1. Study design

This was a pre-planned secondary analysis of Vitamin D for the Immune System in Cystic Fibrosis (DISC) trial [36], which is registered at clinicaltrials.gov as NCT01426256. Briefly, the parent DISC trial was a multicenter, double-blind, placebo-controlled, intent-to-treat clinical trial, spanning five Cystic Fibrosis Foundation Therapeutics Development Network Centers: Emory University and Emory University Hospital (Atlanta, GA), Case Western Reserve University and Rainbow Babies and Children’s Hospital (Cleveland, OH), The University of Alabama Hospital at Birmingham (Birmingham, AL), the University of Cincinnati and University of Cincinnati Medical Center (Cincinnati, OH), and the University of Iowa and University of Iowa Hospitals and Clinic (Iowa City, IA). Before trial participation, all study sites received human studies approval from their local Institutional Review Boards. Within 72 hours of hospital admission for an APE of CF, participants were randomized to receive a single dose of vitamin D_3_ (250,000 IU) orally or placebo. At the 3-month study visit, participants randomized to oral vitamin D_3_ took 50,000 IU of vitamin D_3_ orally every 2 weeks, and participants randomized to placebo took matching placebo orally every 2 weeks. Participants continued their regular vitamin D supplementation if the amount was not greater than 2000 IU daily as recommended by their respective healthcare providers.

### 2.2. Trial eligibility

As previously reported, participants were eligible for the DISC trial if they were 16 years or older, admitted to the hospital for treatment of an APE of CF and enrolled into the study within 72 hours of admission, able to tolerate oral medication, and expected to survive hospital admission [37]. Participants were ineligible if they were unable to provide informed consent, their serum total 25(OH)D concentration was <10 ng/mL or >55 ng/mL within the previous 12 months, had vitamin D intake >2000 IU daily or >10,000 IU vitamin D bolus within the past 60 days, had plans for pregnancy in the next 12 months, had conditions that could be exacerbated by vitamin D such as current hypercalcemia, had conditions that affected vitamin D metabolism such as chronic kidney disease or nephrolithiasis with active symptoms, had conditions that affect survival (such as a history of organ transplantation, HIV/AIDS, illicit drug use, etc.), took oral or intravenous glucocorticoids in the past month, forced expiratory volume in 1-second present predicted (FEV1%) <20%, current hepatic dysfunction with total bilirubin >2.5 mg/dL with direct bilirubin >1.0mg/dL, current use of cytotoxic or immunosuppressive drugs, or were enrolled in any other interventional trial. The primary endpoints of the DISC trial were serial lung function, time to next pulmonary exacerbation, 12-month mortality, and serial plasma cathelicidin concentrations.

In this secondary analysis, participants were excluded if there was no available data on HbA1c or fasting blood glucose level at baseline or the 12-month visit. This was a pre-planned secondary analysis of the DISC parent trial. However, at the initiation of the parent study (2011), studies investigating this topic had not yet been conducted.

### 2.3. Blood Analysis

Routine blood work was collected at the baseline visit during an APE and at the 12-month visit after APE recovery. Values for HbA1c and fasting blood glucose levels were measured by standard hospital methods, and the values were extracted from the electronic medical record.

### 2.4. Statistical analysis

Demographics, HbA1c, and fasting blood glucose levels of participants were reported by mean and their respective standard deviation. P-values for continuous variables were calculated using a two-sample t-test assuming normal distribution and equal variances with significance set at 0.05. A p-value was calculated to detect differences in change in HbA1c and fasting blood glucose between the two groups. Categorical variables were listed as counts and their respective percentages.

## 3. Results

### 3.1. Study participants

Ninety-one participants were enrolled in the parent DISC trial, and 49 participants were eligible for this secondary analysis on glycemic control (Figure 1). The baseline demographic characteristics between the vitamin D_3_ and placebo groups were similar for this sub-group (Table 1). Participants randomized to vitamin D_3_ or placebo had similar baseline serum 25(OH)D, HbA1c, and fasting blood glucose levels (Table 1).

**Figure 1.**
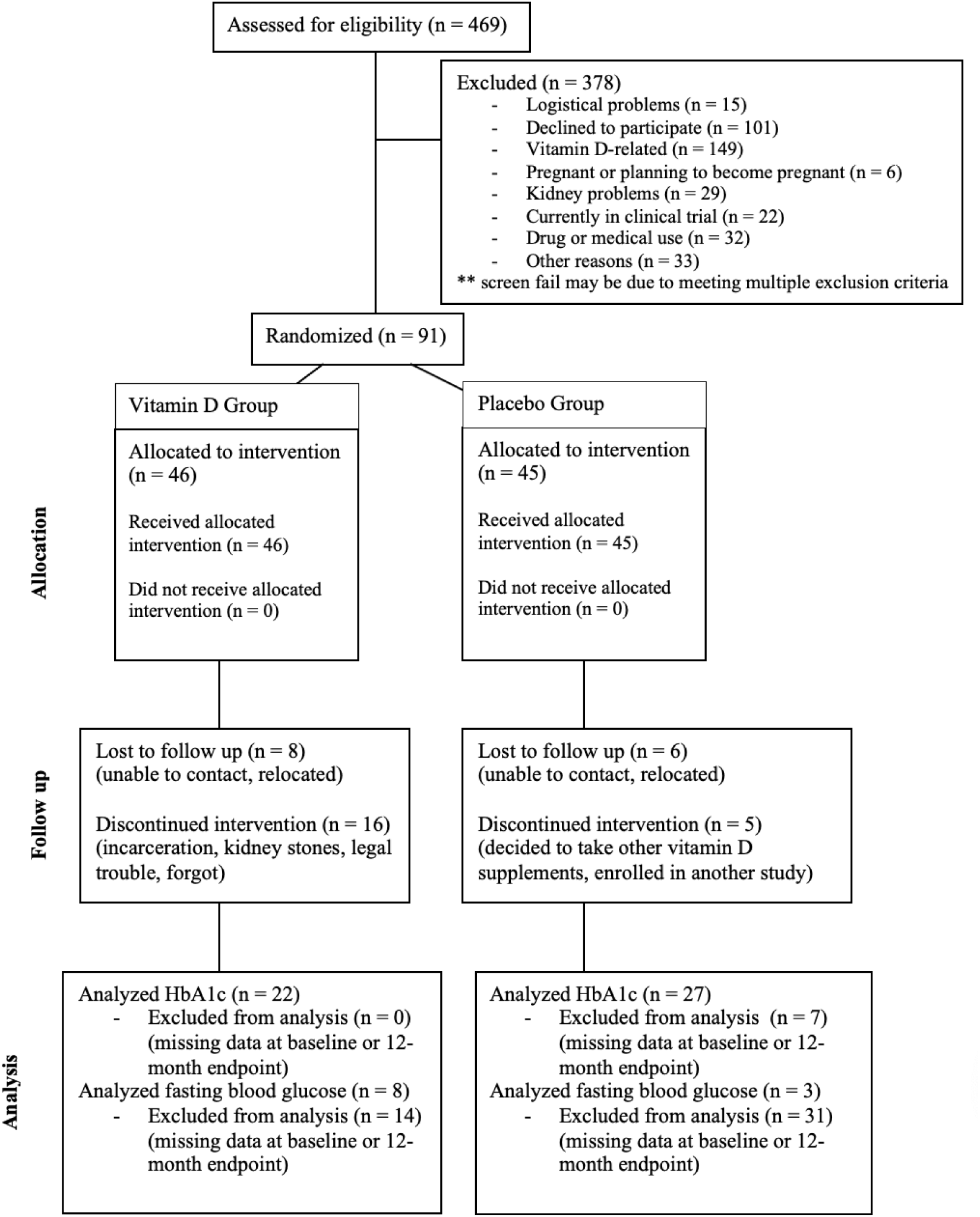
CONSORT diagram of completed DISC study with secondary endpoints.

**Table 1.** Baseline demographics of participants.

|  | **Total (n = 49)** | **Vitamin D<sub>3</sub> (n = 22)** | **Placebo (n = 27)** | **P-value** |
| --- | --- | --- | --- | --- |
| Age (years) | 29.2 ± 8.1 | 29.1 ± 8.0 | 29.3 ± 8.3 | 0.5 |
| BMI (kg/m<sup>2</sup>) | 21.4 ± 4.0 | 21.8 ± 4.6 | 21.0 ± 3.5 | 0.2 |
| Gender (%) |  |  |  |  |
| - Male | 29 (58%) | 11 (47.8%) | 18 (66.7%) |  |
| - Female | 21 (42%) | 12 (52.2%) | 9 (33.3%) |  |
| Race (%) |  |  |  |  |
| - Caucasian | 45 (90%) | 20 (87.0%) | 25 (92.6%) |  |
| - African American | 5 (10%) | 3 (13.0%) | 2 (7.4%) |  |
| CFTR mutation |  |  |  |  |
| - Homozygous for f508del | 45 (90%) | 20 (87.0%) | 25 (92.6%) |  |
| - No copies of f508del | 2 (4%) | 2 (8.7%) | 0 (0%) |  |
| - Unknown | 3 (6%) | 1 (4.3%) | 2 (7.4%) |  |
| Baseline 25(OH)D (ng/mL) | 26.8 ± 9.8 | 28.8 ± 10.8 | 25.2 ± 8.8 | 0.1 |
| Baseline HbA1c (%) | 6.3 ± 1.4 | 6.7 ± 1.8 | 6.1 ± 1.0 | 0.1 |
| Baseline fasting blood glucose level (mg/dL) | 100 ± 29.7<br>(n = 11) | 105.1 ± 33.7<br>(n = 8) | 87.3 ± 8.5<br>(n = 3) | 0.4 |

### 3.2. Change in HbA1c after administration of a bolus dose of vitamin D_3_ or placebo

The mean change in HbA1c for all enrolled study participants was -0.055% ± 0.66. The mean change in HbA1c over 12 months for study participants was not statistically significantly different by treatment group (Table 2). Participants treated with the vitamin D_3_ bolus (n = 22) had a mean HbA1c change of -0.0318% ± 0.949 vs participants treated with placebo (n = 27) had a change of -0.0740% ± 0.290 (p = 0.827) (Figure 2).

**Figure 2.**
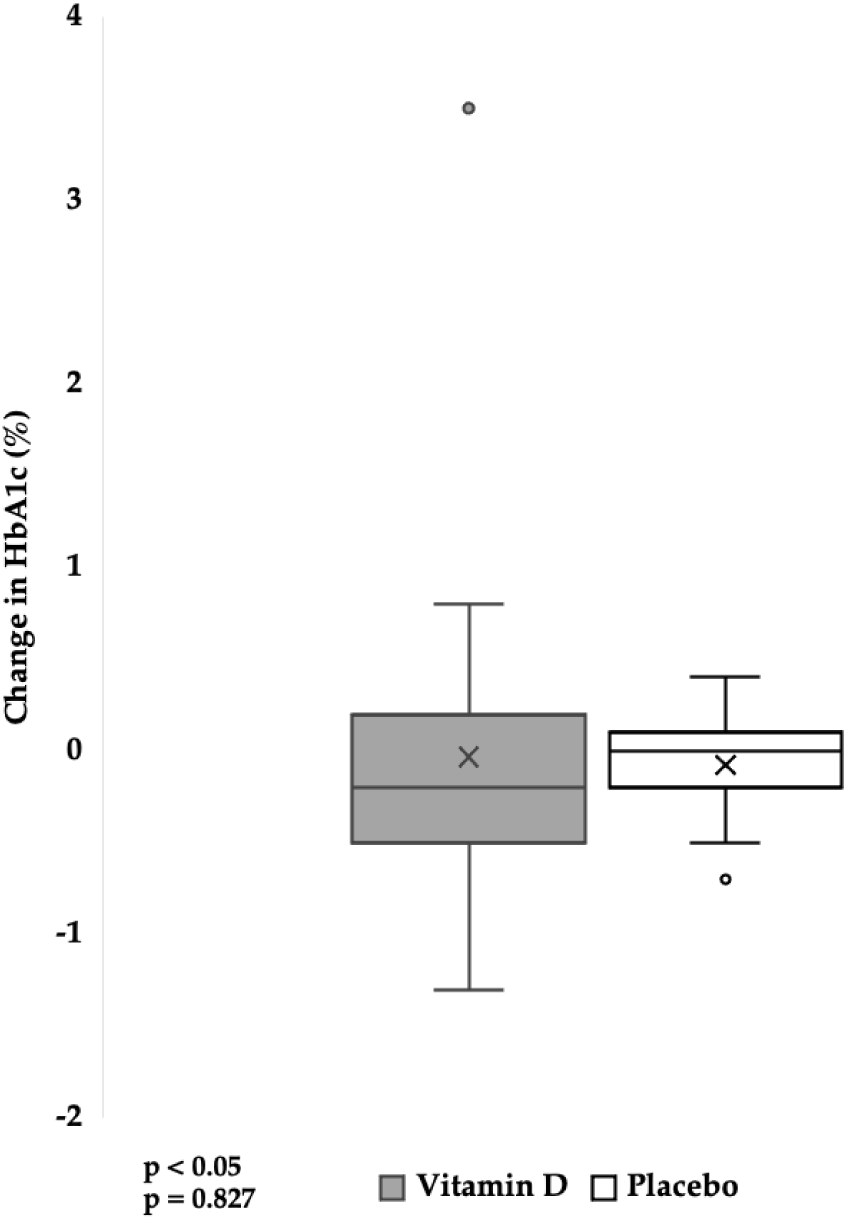
Twelve-month change in HbA1c in study participants with an APE of CF randomized to vitamin D_3_ or placebo.

**Table 2.** Baseline to month 12 change in HbA1c and fasting blood glucose levels in participants randomized to vitamin D_3_ or placebo.

|  | Vitamin D<sub>3</sub> (n = 22) | Placebo (n = 27) | P-value |
| --- | --- | --- | --- |
| Δ HbA1c (%) | -0.03 ± 0.9 | -0.07 ± 0.3 | 0.8 |
| Δ Fasting blood glucose level<br>(mg/dL) | 0.8 ± 39.2 (n = 8) | 15.7 ± 3.8 (n = 3) | 0.5 |

### 3.3. Change in fasting blood glucose after administration of a bolus dose of vitamin D_3_ or placebo

The mean change in fasting blood glucose for all eligible participants was 4.82 mg/dL ± 33.57 (Table 2). The mean change in fasting blood glucose over 12 months for participants was not significantly different by treatment group. Participants randomized to vitamin D_3_ bolus (n = 8) had a mean fasting blood glucose change of 0.75 mg/dL ± 39.19 vs participants treated with placebo (n = 3) had a change of 15.67 mg/dL ± 3.79 (p = 0.54) (Figure 3).

**Figure 3.**
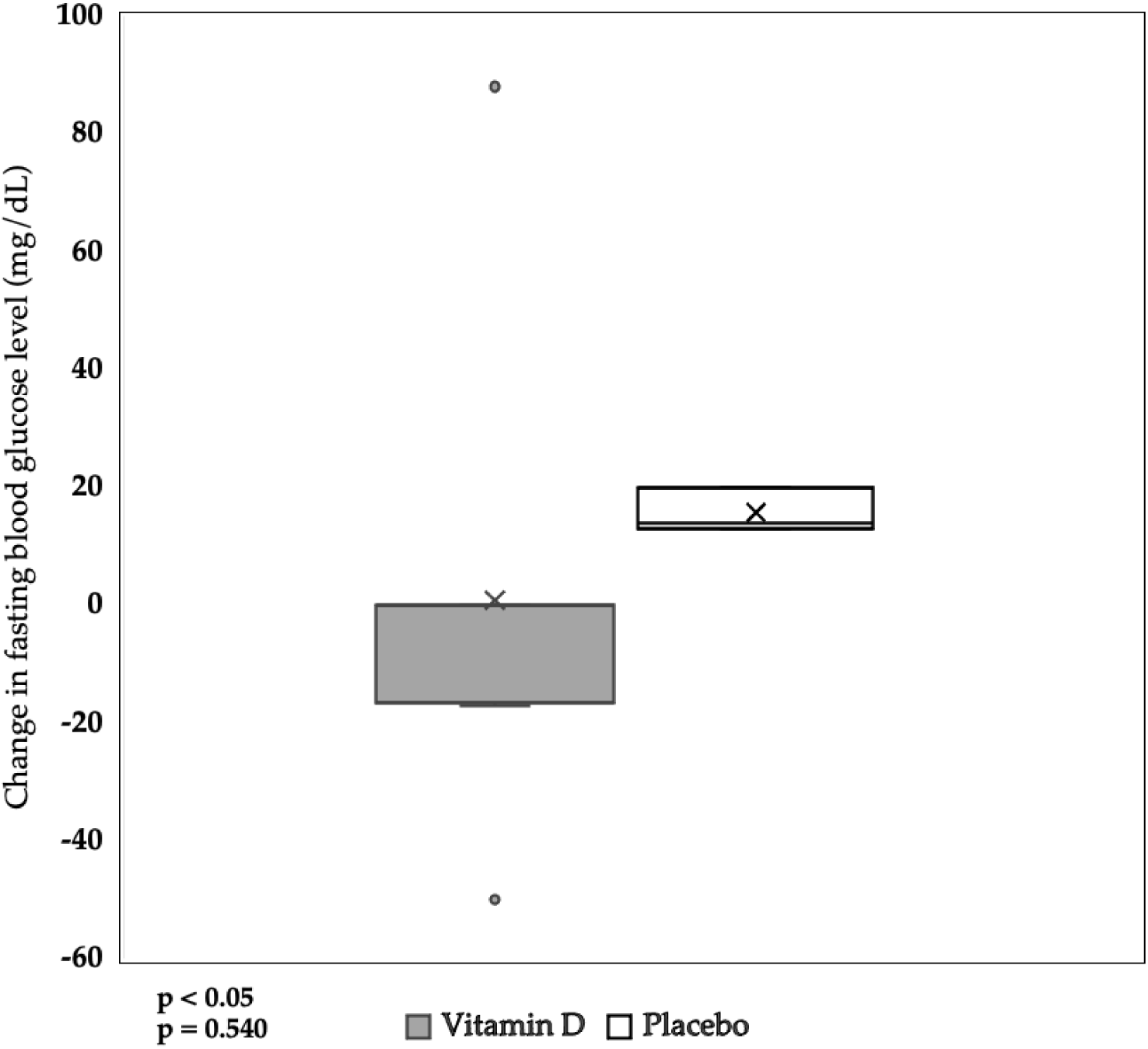
Twelve-month change in fasting blood glucose in study participants with an APE of CF randomized to vitamin D_3_ or placebo.

### 3.4. Baseline to 12-month change in glucose tolerance rating and CFRD status

Glucose tolerance was also measured in 9 of the 50 participants, using the following indicators: 1 - Normal glucose tolerance, 2 - Impaired fasting glucose, 3 - Impaired glucose tolerance, 4 - CFRD with fasting hyperglycemia, 5 - CFRD without fasting hyperglycemia, 6 - Unknown or indeterminate glucose tolerance (Table 3). Of the participants with glucose tolerance recorded (n = 9), 5 participants had normal glycemic control (rating = 1) at baseline and the 12-month follow-up regardless of treatment group. Of those with glucose tolerance measured in the vitamin D group (n = 4), two participants’ glucose tolerance remained the same, one had worsened glycemic control, and one had improved glycemic control. Of participants in the placebo group (n = 5), 4 participants’ glucose tolerance remained the same, and one had improved tolerance. Of the nine participants with glycemic control measured, 2 participants (one in the vitamin D group and one in the placebo group) had CFRD at baseline, and both had improved glycemic control at the endpoint. The participant in the vitamin D group improved from CFRD without fasting hyperglycemia (rating = 5) to only impaired glucose tolerance (rating = 3). The participant in the placebo group improved from CFRD with fasting hyperglycemia (rating = 4) to normal glucose tolerance (rating = 1). Within this nine-patient subgroup, all participants, except one, had stable glucose tolerance (same rating at both time points) (n = 6) or improved glucose tolerance (n = 2) from baseline to the 12-month endpoint regardless of the treatment group.

**Table 3.** Baseline and month 12 glucose tolerance rating with the following scale: [1 - Normal glucose tolerance, 2 - Impaired fasting glucose, 3 - Impaired glucose tolerance, 4 - CFRD with fasting hyperglycemia, 5 - CFRD without fasting hyperglycemia, 6 - Unknown or indeterminate glucose tolerance].

| **Vitamin D group (n = 4)** | **Baseline rating** | **Month 12 rating** |
| --- | --- | --- |
| Patient 1 | 5 | 3 |
| Patient 2 | 1 | 3 |
| Patient 3 | 3 | 3 |
| Patient 4 | 1 | 1 |

| **Placebo group (n = 5)** | **Baseline rating** | **Month 12 rating** |
| --- | --- | --- |
| Patient 1 | 1 | 1 |
| Patient 2 | 4 | 1 |
| Patient 3 | 1 | 1 |
| Patient 4 | 1 | 1 |
| Patient 5 | 1 | 1 |

## 4. Discussion

The purpose of this secondary analysis was to determine the role of a single high-bolus dose and maintenance vitamin D dosing of vitamin D_3_ on glycemic control in participants with CF after an APE. Based on the understanding that a critical illness can commonly cause an increase in insulin resistance and hyperglycemia, we intended for this secondary analysis to examine if there were any therapeutic effects of vitamin D supplementation to counteract the likely decline in glycemic control following an APE. We found that administration of a single vitamin D_3_ bolus in participants with CF at the time of hospital admission for an APE followed by bi-weekly bolus doses of vitamin D for one year did not result in significant changes in HbA1c and fasting blood glucose levels compared to placebo. Despite this, all participants with glucose tolerance measured, except one, had stable glucose tolerance (same rating at baseline and month 12) (n = 6) or improved glucose tolerance (n = 2) regardless of the treatment group they were assigned. Additionally, participants randomized to vitamin D_3_ showed fasting blood glucose levels increased less than those randomized to placebo, although, these results were underpowered. The increase in fasting blood glucose levels for both groups could be considered predictable given the history of an APE, and therefore, vitamin D supplementation may have mitigated the negative effects on glycemic control that were caused by the APE. Our findings suggest that the administration of a vitamin D_3_ bolus dose may not be beneficial for long-term glycemic control in participants with CF or that the duration and/or dosage of vitamin D administration were not adequate to detect a significant difference.

Our secondary analysis determined that vitamin D supplementation after an APE did not induce a significant improvement in glycemic control in patients with CF. Therefore, it may not be beneficial for patients with CF, who are not vitamin D deficient, to take supplemental vitamin D. However, it should be considered that during an APE, patients with CF very commonly develop hyperglycemia, which seems to not improve in the long-term [34]; we may have not detected significant changes due to this consideration. Because vitamin D may not have a significant therapeutic effect on glycemic control, it is important to consider alternatives that may be more beneficial, given that hyperglycemia and lung function decline seem to be associated. A meta-analysis of randomized controlled trials (RCTs) on type 2 diabetes treatments, specifically sodium glucose cotransport-2 (SGLT-2) inhibitors and glucagon-like peptide-1 (GLP-1) receptor agonists, indicated improved health-related quality of life, reduced all-cause mortality, improved end-stage kidney disease, and lowered risk of hospitalization for heart failure; these two drug classes are specifically beneficial to adults with elevated risk for cardiovascular and kidney complications [38]. However, SGLT-2 inhibitors are associated with genitourinary infections, ketoacidosis, acute renal injury, etc. [39], and GLP-1 receptor agonists are associated with pre-renal acute kidney injury and various mild gastrointestinal symptoms [40]. An umbrella review of Mendelian randomization (MR) studies and RCTs indicated low vitamin D was a causal risk factor for type 2 diabetes [41]. The meta-analysis concluded vitamin D supplementation may have some therapeutic and long-term preventative effects on chronic diseases (such as type 2 diabetes, hypertension, Alzheimer’s disease, and schizophrenia) and decrease all-cause mortality; however, the current implementation, which utilizes vitamin D as an intervention for the diseases, might not be efficient, thus, more effective strategies are needed to improve the outcomes of vitamin D supplementation as an intervention for chronic diseases [41]. Adults with higher 25(OH)D levels (genetically predicated and lifelong) are at higher risk for kidney stone disease, partially via elevated calcium levels [42]. In comparing the risks and benefits of vitamin D supplementation vs. drugs designed for type 2 diabetes, vitamin D is less likely to induce severe side effects (unless there is prior genetic predisposition). Therefore, it would be extremely beneficial to elucidate a highly efficient and effective method utilizing vitamin D as an interventional therapeutic.

In the context of CF, hyperglycemia is highly prevalent during pulmonary exacerbations [34]. Animal models show that increased glucose concentration within the airways promotes the growth of bacteria and negatively alters immune function, reducing phagocytosis of neutrophils and chemokinesis [13, 43, 44], possibly leading to more frequent APEs and worsened recovery. Upon hospital admission for an APE, a study showed 8/9 pediatric patients with CF had hyperglycemia (2 CFRD, 6 abnormal oral glucose tolerance test (OGTT)), and 2 weeks later, 5/8 had hyperglycemia (2 CFRD, 3 abnormal OGTT); at follow-up (3.8-6.3 months), glucose tolerance declined after the initial improvement [34]. It is possible that hyperglycemia initially improved after antibiotic treatment for the pulmonary exacerbation due to reduced inflammation, as inflammation is associated with increased insulin resistance and decreased secretion of insulin; however, long-term effects of exacerbation-associated inflammation or untreated hyperglycemia may have contributed to damaging or toxic effects on beta cells, reducing beta cell function and insulin secretion [34]. Without treatment for hyperglycemia after an APE, there seems to be glycemic control improvement in the short term but a decline in the long term.

Further, morbidity and acquired infections during hospitalization significantly decrease after treating hyperglycemia [44]. Therefore, based on previous studies showing some therapeutic promise of vitamin D supplementation, we proposed vitamin D may be able to treat hyperglycemia to improve the quality of life of patients with CF after an APE.

Several studies show that vitamin D is needed for optimal glucose metabolism and homeostasis [26, 45, 46]. Vitamin D intake is believed to decrease the prevalence of type 2 diabetes through stimulation of gene expression of insulin receptors and improving transport of glucose from the intestine [47, 48]. A study in human promonocytic cells showed that insulin receptor capacity and insulin-stimulated glucose transport increased when activated by 25(OH)_2_D_3_ [25, 48, 49]. Vitamin D supplementation was seen to improve glycemic control in only participants with vitamin D deficiency and had no therapeutic effects for participants already within the normal range of 25(OH)D [50]. These previous studies suggest that vitamin D may be important for glucose metabolism in people who have vitamin D deficiency and who are at risk for diabetes. This may be the reason for the lack of therapeutic effects in this secondary analysis.

However, some prior research supports our findings that vitamin D supplementation does not improve glycemic control. A study showed that vitamin D_3_ supplementation normalizes 25(OH)D concentration in patients with diabetes but may not have long-term effects on glycemic control [51]. The large-scale Vitamin D and Type 2 Diabetes (D2d) study (n = 2423) concluded that two-year vitamin D supplementation, 4,000 IU daily, in patients with vitamin D deficiency did not result in significantly lower risk of diabetes as compared to placebo, despite differences in 25(OH)D_3_ at month 24 (54.3 ng/mL in vitamin D group vs 28.8 ng/mL in placebo group) [25]. Further, a systematic review of 13 randomized controlled trials on the role of vitamin D supplementation in the prevention of type 2 diabetes indicated that vitamin D supplementation had no statistically significant impact on the incidence of type 2 diabetes in six of the seven trials that identified type 2 diabetes incidence as a research outcome [52–65]. Additionally, 10 of the trials analyzed the impact of vitamin D supplementation on insulin resistance using the Homeostatic Model Assessment for Insulin resistance (HOMA-IR). In six of the trials, patients who received vitamin D supplementation had lowered and improved insulin resistance while the four remaining trials had conflicting results [52]. However, these trials ranged from 12 weeks to 60 months, in which vitamin D dosage, vitamin D-deficient status, diabetes status, and body mass index (BMI) varied widely, possibly contributing to conflicting results on the efficacy of vitamin D [52]. In a separate review, where over 15 RCTs were reviewed, vitamin D supplementation did not improve HbA1c, insulin resistance, or fasting blood glucose [35].

However, in four of the trials, prediabetic subjects who received vitamin D supplementation had improved fasting plasma glucose [52]. Similarly, Farahmand et. al. found that vitamin D supplementation was associated with reduced HbA1c when participants were vitamin D deficient at baseline [66]. Thus, prior findings have indicated conflicting results regarding the therapeutic effects of vitamin D on glycemic control, especially regarding optimal dosage and duration. It remains unresolved whether specific populations, such as those with prediabetes, may benefit from vitamin D supplementation.

Study limitations include the number of participants eligible within the secondary study, the timing of vitamin D_3_ intervention, the severity of the APE, baseline vitamin D status, and only one end timepoint (12-month) used for comparison to baseline. Future studies may find it useful to examine glycemic biomarkers at additional time points. As a secondary analysis, the sample size may not have been large enough to conclude statistically significant changes in glycemic control. However, this data provides some initial impressions of the effect of this intervention on glycemic control after an APE and may inform the study design of future studies. The participants involved in the study were admitted following an APE, in which the timing of the bolus dose of vitamin D_3_ may not be appropriate for improvement in glycemic control. After critical illness, such as an APE, it is common for hyperglycemia and insulin resistance to develop [33], possibly explaining the rise in HbA1c despite vitamin D supplementation. In a study conducted in 9 pediatric patients following CF exacerbation, hyperglycemia was highly prevalent, which seemed to improve after treatment for the exacerbation and worsen later on when insulin secretion decreased as well [34]. CFRD development has been associated with worsened lung function and disease, and insulin therapy has been seen to improve these symptoms [11]. A retrospective study in children showed recovery from exacerbations can be altered by untreated hyperglycemia [12]. Following hospital admission, many participants were recovering from the effects of APE and still receiving antibiotics. Prolonged use of antibiotics has been associated with a higher risk of developing diabetes [67]. Additionally, at baseline, participants in both groups had a mean 25(OH)D_3_ level considered adequate (>20 ng/mL). In our analysis, at baseline in the vitamin D group, participants’ 25(OH)D was 28.8 ng/mL ± 10.8, and participants in the placebo group’s 25(OH)D was 25.2 ng/mL ± 8.8. Therefore, on average, participants in both groups were not vitamin D deficient at baseline. Previous studies show that vitamin D_3_ supplementation was beneficial in instances where patients were vitamin D deficient (25(OH)D <20 ng/mL) [50]. In future studies, it may be beneficial to measure HbA1c and fasting blood glucose at more time points to evaluate both the short-term and long-term effects of an APE on hyperglycemia as well as the effect of vitamin D on glycemic control at different time points during recovery from the APE. This secondary analysis provides initial impressions of the effect of vitamin D supplementation on an APE of CF and may inform future study designs based on these limitations.

In conclusion, high-dose bolus vitamin D_3_ treatment upon hospital admission for an APE, followed by maintenance vitamin D dosing, did not improve glycemic control after 1 year in adults with CF. Although there are many conflicting studies regarding the effect of vitamin D supplementation on hyperglycemia, the results from our secondary analysis of the DISC study support some prior findings that vitamin D supplementation may not improve glycemic control despite its possible effect on insulin resistance. However, the added nuance of previously documented hyperglycemia and insulin resistance during and after an APE should be considered, as vitamin D may have had some therapeutic effect that was not detected or significant. More adults with CF are developing CFRD due to increasing rates of survival and increased incidence of overweight and obesity. The findings from this secondary analysis suggest that high-dose bolus vitamin D_3_ administration (during hospital admission of an APE of CF) and patient population (not deficient in vitamin D_3_ and/or prior glucose tolerance status) did not impact glycemic control. Future studies may elucidate whether poor glycemic control results in declined FEV1 recovery or if biomarkers for poor glycemic control are indications of worsened disease. Further research with rigorous evaluation of blood glucose control with continuous glucose monitoring and more frequent measurements of HbA1C over time is required to investigate the impact of vitamin D_3_ on patients with CF after an APE, especially those with vitamin D deficiency and at high risk for diabetes.

## Data Availability

All data produced in the present study are available upon reasonable request to the authors.

## Author Contributions

The authors’ contributions were as follows – VT and JAA: designed the study and obtained funding; VT, JAA, TRZ, and WRH: performed and supervised the conduct of the trial and revised the manuscript; AS and VT: performed secondary analysis, verified, interpreted the data, and revised the manuscript; AS performed statistically analyses and drafted the manuscript.

## Acknowledgments

The study investigators would like to acknowledge the participants and study team in the Vitamin D for Enhancing the Immune System in Cystic Fibrosis clinical trial.

